# High genetic diversity of *Histoplasma* in the Amazon basin, 2006-2017

**DOI:** 10.1101/2025.04.01.25324933

**Authors:** Tani Ly, Marcus de Melo Teixeira, Gaston Jofre-Rodriguez, Denis Blanchet, Sigrid Mc Donald, Primavera Alvarado, Silvia Helena Marques da Silva, Victoria E Sepúlveda, Qandeel Zeb, Stephen Vreden, Antoine Adenis, Francisco Yegres, Magalie Demar, Maria José Serna Buitrago, Bridget Barker, Mathieu Nacher, Daniel R. Matute

**Affiliations:** UMR TBIP, Université de Guyane, 97300 Cayenne, French Guiana; CIC INSERM1424, Institut Santé des Populations en Amazonie, Centre Hospitalier de Cayenne, 97306, Cayenne, French Guiana; Faculty of Medicine, University of Brasília, Brasília-DF, Brazil; Department of Biology, Virginia Commonwealth University; Laboratoire de Parasitologie-Mycologie, Centre Hospitalier de Cayenne, 97300 Cayenne, French Guiana; Department of Medical Microbiology, University of Amsterdam UMC, Amsterdam, Netherlands. Evandro Chagas Institute, Bacteriology and Mycology Section, Mycology Laboratory, Ananindeua, Pará, Brazil; Laboratorio de Micología. Instituto de Biomedicina “Dr. Jacinto Convit”. Caracas, Venezuela; Facultad de Medicina, Universidad Central de Venezuela; Evandro Chagas Institute, Bacteriology and Mycology Section, Mycology Laboratory, Ananindeua, Pará, Brazil; Biology Department, University of North Carolina, Chapel Hill, NC, USA; Stichting Wetenschappelijk Onderzoek Suriname, Paramaribo, Suriname. Academisch Ziekenhuis Paramaribo, RRP8+PQ7, Flustraat, Paramaribo, Suriname; CIC INSERM1424, Institut Santé des Populations en Amazonie, Centre Hospitalier de Cayenne, 97300, Cayenne, French Guiana; Centro de Investigaciones Biomédicas, Universidad Nacional Experimental Francisco de Miranda, Coro, Estado Falcón, Venezuela; Laboratoire EA3593 Ecosystemes amazoniens et pathologies tropicales, Université de Guyane, 97300 Cayenne, French Guiana; Laboratoire de Parasitologie-Mycologie, Centre Hospitalier de Cayenne, 97300 Cayenne, French Guiana; Centro Nacional de Microbiología, Instituto de Salud Carlos III, 28220 Majadahonda, Madrid, Spain; CIBERINFEC, ISCIII -CIBER de Enfermedades Infecciosas, Instituto de Salud Carlos III; Pathogen and Microbiome Institute, Northern Arizona University, Flagstaff-AZ, USA

## Abstract

Genome sequencing has revealed that *Histoplasma*, the etiological agent of histoplasmosis is composed of several phylogenetic species. Nonetheless, the genetic diversity of the pathogen remains largely unknown, especially in the tropics. We sequenced the genome for 91 *Histoplasma* isolates from the Amazon basin, and used phylogenomics, and population genetic evidence to measure the genetic variation of the genus in South America. We report a previously unidentified clade of *Histoplasma* endemic to the Amazon basin. The lineage is widespread across the continent and contains five lineages that are sufficiently differentiated to be considered phylogenetic species. We find that the geographic range of these lineages is largely but not completely overlapping. Finally, we find that the patient median age and sex ratio differs among species suggesting differences in the epidemiology of histoplasmosis caused by each *Histoplasma* lineage.

## INTRODUCTION

Fungal diseases have a large and negative effect on human well-being (1). Among mycoses, histoplasmosis is one of the most common pulmonary diseases in the world (2). In immunocompetent hosts, most cases of histoplasmosis are mildly symptomatic and self resolutive. In these patients the disease is not of mandatory report and is often undiagnosed, if not misdiagnosed. Histoplasmosis is of paramount importance among immunosuppressed patients. In some areas, up to 25% of the HIV-positive population develops histoplasmosis which frequently turns fatal (3). Approximately half a million people are diagnosed with histoplasmosis every year and close to 100,000 people develop a progressive disseminated disease (4). Among persons with advanced HIV (i.e.,CD4 cell count <200 cells/mm^4^), the disease has a case-fatality rate between <5% to 50% when treated (5, 6) and close to 100% if not treated (6).

The etiological agent of histoplasmosis, *Histoplasma* sp., is a cosmopolitan fungus detected in all continents (7) including Antarctica (8). Histoplasmin—the main *Histoplasma* antigen—skin testing has revealed that the fungus has a large geographic range (9). Nuclear gene genealogies and a global sample of *Histoplasma* strains from eight countries revealed the existence of at least seven phylogenetic species (7,10), monophyletic groups that are reciprocally monophyletic and isolated from each other. Genome sequencing has confirmed the existence of differentiated phylogenetic species within *Histoplasma* (11,12). These assessments have been limited as the sampling for *Histoplasma* has been heavily biased towards North America and because few samples from other locations have been fully sequenced.

Nonetheless, histoplasmosis is rampant throughout the Americas. South American patients suffer from disseminated histoplasmosis at one of the highest incidences in the world (1.5 cases per 100 person-years in persons living with HIV, 13). Histoplasmin surveys have detected multiple localized foci of high skin reactivity to *Histoplasma* (14). Preliminary genetic analysis suggested that isolates from South America are genetically diverse (10,15). More recent approaches have used genomic data and revealed the existence of a phylogenetic species endemic to Rio de Janeiro in southern Brazil (16). Yet, the *Histoplasma* isolates from South America, a continent that has been hypothesized as a reservoir of diversity for the genus (7,10), remain largely uncharacterized genetically. A systematic study of the genome-wide diversity of *Histoplasma* across the Americas is sorely needed. In this report, we use whole genome sequences to measure the genetic diversity of *Histoplasma* across America. We sequenced 91 genomes of South and Central American isolates, used data from previous sequencing efforts, generated the largest phylogenetic assessment for this pathogen to date (187 genomes), and studied the extent of divergence within American *Histoplasma*. We identified five lineages that meet the criteria for being classified as phylogenetic species and compared the epidemiology of histoplasmosis caused by each lineage. Our findings reveal significant differentiation among these lineages, as well as variations in the epidemiology of histoplasmosis associated with each one. Our results highlight the need of integrating epidemiological, evolutionary, and clinical studies to understand the importance of genetic divergence in fungal pathogens.

## MATERIALS AND METHODS

### Fungal isolates

We obtained cultures from patients with clinically-defined histoplasmosis between 2006 and 2017 by subculturing samples on Sabouraud agar with chloramphenicol, gentamycin, and actidione (Bio-Rad Laboratories, Hercules, CA). Table S1 lists the collection site, sex, age and the type of sampling and disease for each patient. After obtaining pure mycelial cultures, each clinical isolate was subcultured on the same media type to obtain enough fungal biomass for DNA extraction (at least 500mg). DNA extraction protocols are summarized in the Appendix.

### Reference genome for *Histoplasma* mz5-like

We assembled a *de novo* genome for the *Histoplasma suramericanum* strain mz5, an isolate originally collected in Colombia, using Oxford Nanopore technologies (ONT) long-reads. We obtained a total of 231,650 reads, with an average length of 4,681.3 bp (SRA accession number: TBD). The mean coverage from our reads was 31.13. We used Flye (17) to assemble the reads and three runs of Racon (18) and Medaka (19) to iteratively polish the assembly. We used Pilon (20) for indel corrections four times using the FASTQs files. To assess the quality and completeness of our resulting assembly, we used Quast (21) and BUSCO (22) with the fungi and Eurotiomycetes OrthoDB V10 databases (23). The assembly had 26 contigs; 95.92% of the genome was assembled in the ten largest contigs and we focused on those for the phylogenetic analysis.

### Isolate resequencing

To prepare genomic libraries, we used KAPA library preparation kits for Illumina NGS sequencing (Kapa Biosystems) and 1μg of purified DNA per isolate. Next, we indexed the libraries using unique 8-bp nucleotide identifiers. We evaluated the concentration of each library with a Kapa library quantification kit (Kapa Biosystems, Wilmington, MA) on a 7900HT Instrument (Life Technologies). We sequenced the libraries to a read length of 100 bp using v3 or v4 chemistries on an Illumina HiSeq 2500 instrument or to 150bp using v2 chemistry on an Illumina NextSeq platform (Illumina, San Diego, CA), both on a high output mode (paired-end). The coverage and accession numbers for each isolate is shown in Table S2.

### Previously published data

To compare South American isolates of *Histoplasma* with isolates from other locations, we used 30 previously sequenced genomes deposited by (12). We also used 16 genomes from a *Histoplasma* lineage endemic to India (24) and fifty genomes from Rio de Janeiro (16). To root the phylogenetic trees (see below), we used *Paracoccidioides* genomes from two different species (*P. restrepiensis, N* = 3; *P. brasiliensis sensu stricto N* = 2; 25), *Blastomyces* (*N* = 5; 26), *Emmonsia crescens* (*N* = 2; 27) and *Emergomyces pasteurianus* (*N =* 1; 27). The SRA accession numbers of all these genomes are listed in Table S3.

### Read mapping

We used one of the samples from the newly identified Latin American lineage (see Results) to produce a new reference genome for the group (strain mz5). This reference genome has a total length of 34,827,701 bp, with 26 contigs, an N50 of 5161774, and a BUSCO completeness assessment of 98.6% complete Eurotiomycetes orthologs, and 98.5% complete fungi orthologs (22). We mapped 200 short-red sequenced samples (91 newly sequenced genomes) to the reference. This brings the number of *Histoplasma* genomes from the Americas to 113. Protocols to call variants are listed in the Appendix.

### Phylogenetic analysis

To study the genealogical relationships among *Histoplasma* isolates, we converted our multisample VCF file into a concatenated genome-wide alignment in Phylip format using the Python script *vcf2phylip* (28). We then extracted the 10 largest contigs from our multisample VCF using *bcftools* (29). Then, we built Maximum Likelihood (ML) trees from each of the 10 largest contigs and the genome-wide alignment using IQ-TREE 2 (30). ModelFinder (31) determined that TVMe+R3 was the best-fitting model of nucleotide substitution for the genome-wide alignment. (Table S4 shows the model fitting for different models of molecular evolution.) To estimate branch support we generated 1,000 tree replicates with an ultrafast bootstrap approximation (32). We used a similar approach to generate genealogies for the largest ten supercontigs in the nuclear genome. We compared these trees using a Robertson-Foulds (RF) distance (33) as implemented in the R function *treedist* (library *phangorn,* 34).

Second, we estimated the extent of genealogical concordance for the nuclear genome in two ways. First, we studied whether different genomic windows showed the same genealogy. We split into 325 non-overlapping windows, each of 100 kb, and used IQ-TREE 2 (30) to generate a genealogy from each partition (325 genome-window trees; different partitions gave similar results.) We calculated the concordance factors (CF; 35,36) as the fraction of genealogies concordant with each branch from the species trees. Second, we used gene trees from 3,494 complete Eurotiomycetes orthologs (eurotiomycetes_odb10; 22,23). Our approach to generate the gene trees is identical as described for gene genealogies. Lineages that showed reciprocal monophyly, and had high levels of concordance among supercontigs, were treated as potential phylogenetic species for further analyses (following 37).

### Genetic diversity and differentiation

We used population genetics approaches to study the partition of genetic variation in *Histoplasma*. We estimated the magnitude of genetic variation (π) within each lineage of *Histoplasma* (as identified by the concatenated phylogenetic tree and the concordance analysis, see immediately above) and compared these values to the magnitude of pairwise divergence between species (Dxy). Instances of advanced speciation show a significantly higher Dxy between the two focal groups than the π value of either group (38). We used *Pixy* (39) for all calculations. To compare the values of π in each of these lineages with the pairwise D_xy_, we used an Approximative Two-Sample Fisher-Pitman Permutation Test (R function *oneway_test*, library *coin*, 40).

### Patient characteristics for each *Histoplasma* lineage

Finally, we studied general epidemiological patterns of histoplasmosis caused by each lineage identified in this study. For clinical isolates, we collected age, sex, HIV status and a description of the disease (i.e., whether histoplasmosis was disseminated). Table S1 lists the anonymized clinical data for all isolates included in this study. We did three analyses using this dataset. First, we compared whether the four countries with the largest number of cases (French Guyana, Brazil, Suriname, and Venezuela) had similar proportional representation of the six resident *Histoplasma* phylogenetic species (See Results) using a 2-sample test for equality of proportions with continuity correction (*prop.test* function, R library *stats*; 41). We did six pairwise comparisons and adjusted the P-values using a Bonferroni correction (function *p.adjust*, R library *stats*; 41). We calculated the power of the proportion tests using the function *pwr.2p2n.test* (R library *pwr;* 42).

Second, we studied whether reports of histoplasmosis were equally common in males and females (i.e., whether the sex ratio of histoplasmosis patients was 1:1) across lineages. We used a *X^2^* test using the R function *chisq.test* (library *stats*; 41). We calculated the power of each *X*^2^ test using the function *power.chisq.test* (R library *DescTools*; 43). We only report the comparisons for the two lineages that had a X^2^ power equal or larger than 0.5. We also compared whether the patient sex proportional representation differed among lineages using a linear model (R library *stats;* 41; *glm*: family = "binomial").

Finally, we compared the age of the histoplasmosis patients affected by each of the five lineages. We used a type-III ANOVA (function *Anova*, R library *car*, 44), followed with Tukey Honestly Significant Difference post-hoc pairwise comparisons (function *glht*, library *multcomp*; 45) to identify whether lineages differed from each other.

## RESULTS

Here we report *Histoplasma* collections for 11 years (2006-2017) in French Guiana, Suriname, Brazil, Venezuela, Guyana, Martinique, Nicaragua, and Spain. With these isolates in hand, we used a phylogenetics and population genetics lens to understand the epidemiological patterns of histoplasmosis in the Amazon basin and adjacent areas.

First, we used this genome-wide data to resolve the phylogenetic relationships between *Histoplasma* lineages. A maximum likelihood phylogenetic tree for all samples of *Histoplasma* using concatenated markers revealed the existence of a monophyletic group composed from South and Central American *Histoplasma* (Figure 1A). The clade includes *H. suramericanum*. We refer to this clade as the Latin American *Histoplasma* clade but do not imply that the group does not contain unsampled lineages from outside Latin America. Most previously sequenced samples, notably from North America, some from South America, and India (14,24) form a second monophyletic group which we refer to as the global *Histoplasma* species complex. The global species complex includes eight lineages (seven with more than one isolate): one from Africa, one from India, and six American lineages: *H. ohiense*, *H. mississippiense*, LAm B, *H. capsulatum ss*. -originally thought to be restricted to Central America, and two poorly sampled lineages of clinical provenance (27-14, one isolate; and clinical isolates from Brazil, two isolates). The two latter ones were not previously described but all other lineages have been reported previously (14,24).

**FIGURE 1.**
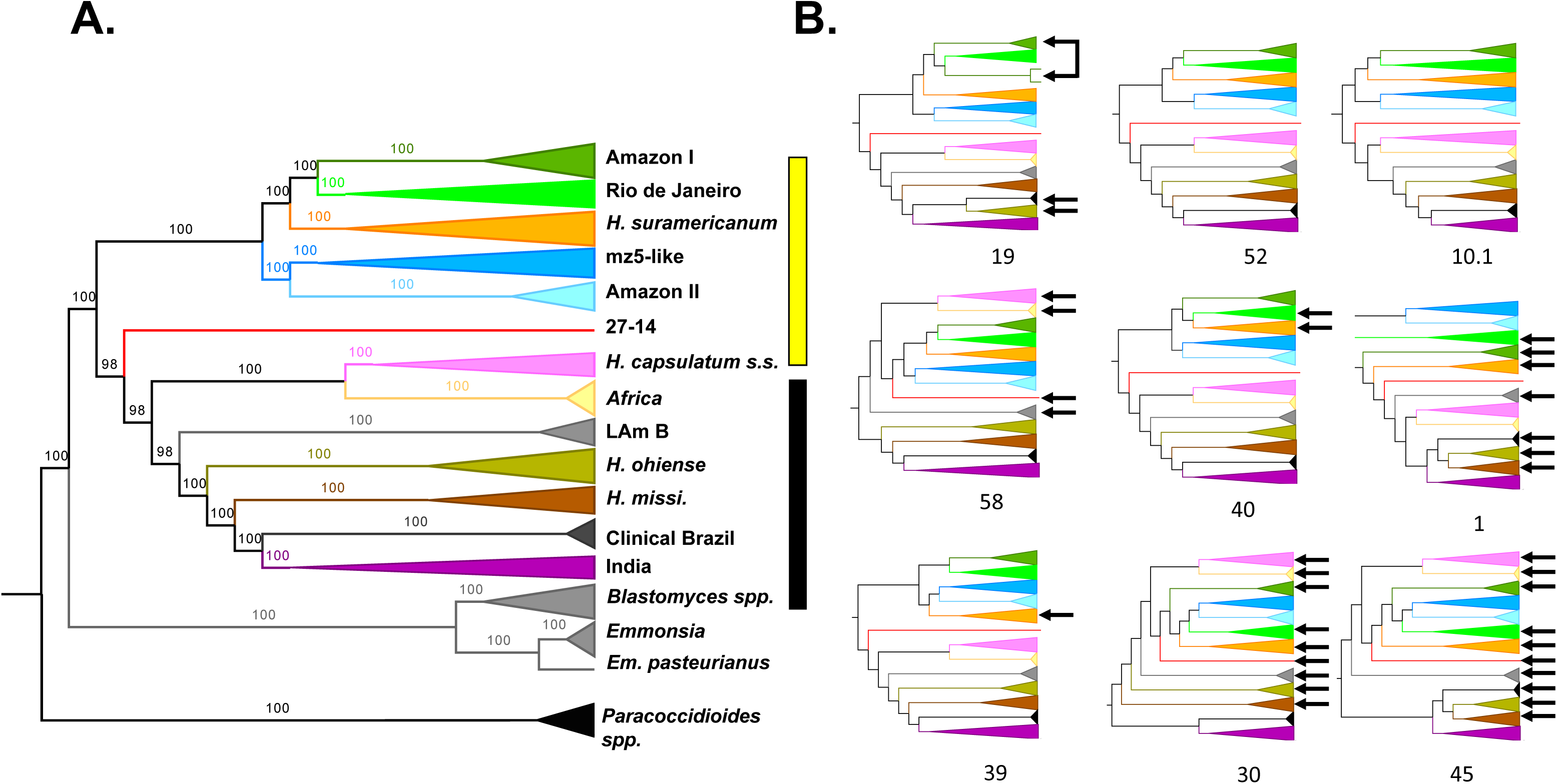
Phylogenetic analyses reveal a large level of genetic diversity of *Histoplasma* in South America. Whole-genome concatenated markers show the existence of at least twelve monophyletic groups. The Latin American clade is marker with a yellow bar; the global clade is marked with a black bar. Figure S1 shows concordance analyses using BUSCO genes (S1) and genomic windows (S2). **B**. The nine largest supercontigs show largely consistent topologies. Arrows show lineages with positions that differ from the inferred species tree. The color scheme is the same as shown in A. Figure S2 shows the RF distances between trees.

The Latin American lineage only includes samples from South and Central America and is highly diverged from most of the previously sequenced samples of *Histoplasma* from around the world. This lineage contains five clades (*H. suramericanum* (11), Amazon I, Amazon II, and RJ (16), and mz5-like). Following a mutation rate similar to other ascomycetes (46), the age of the Latin American lineage is on the order of 3.2 million years (CI= 0.9 MYA). We evaluated whether the five monophyletic groups in the Latin American radiation revealed by the concatenated tree fulfilled the requirements to be considered different phylogenetic species. We tested two additional criteria to assess whether they were differentiated enough in the speciation continuum. First, in cases of advanced speciation, different genome sections show consistent evolutionary trajectories. The five groups appear as monophyletic in the concatenated analyses which is consistent with the possibility of each of these lineages being phylogenetic species. All the clades, apart from the RJ lineage, co-occur in the Amazon basin. We evaluated whether the signal from the concatenated genome was also consistent at a more granular level (Figure 1B). Local ancestry analyses are consistent with the genome-wide results. Concordance between supercontigs was high, but not perfect, and some supercontigs revealed some variations in the inferred phylogenetic relationships (Figure 1B, Figures S1 and S2).

Concordance factors at the genome window level showed that four of the five lineages had moderately high concordance factors (CF; >50%; Figure S1) but that one group, the lineage from Amazon I, showed a low CF (17%) which suggests diverse genetic trajectories along the genome. Notably, this is the same lineage that showed non-monophyly in supercontig 19 (Figure 1B). Figure S2 shows the RF distance between pairs of supercontigs and with the tree inferred from a concatenated alignment (shown in Figure 1A). In general, recent divergences have high bootstrap support and concordance factors, but older splits had lower support (Figures 1, S1, and S2). Despite these genealogical differences, the genome wide and local ancestry results indicated a high level of phylogenetic concordance among different genomic regions.

Second, the genetic distance between isolates of different species tends to be larger than the extent of polymorphism within species (37,47). We evaluated whether the extent of genetic differentiation (Dxy) between the monophyletic lineages was larger than the magnitude of genetic variation within lineages (π). We found that in almost all pairwise assessments (74 out of 78, Table S5), Dxy was larger than the variation in any of the lineages, suggesting advanced divergence in Latin America (Figure 2). The four pairs that were not significant involved lineages with only two isolates, 27-14 in three cases, and ‘clinical isolates from Brazil’ in two cases (one of the pairs is clinical isolates-27-14; Table S5). Notably, there is a weak association between heterozygosity and geographic range size (Spearman’s rank correlation ρ=0.685, *P* = 0.035), indicating the larger ranges might serve as large reservoirs of genetic variation in *Histoplasma* (Figure 3A).The extent of genetic differentiation as revealed by both phylogenetic and population genetics approaches suggests that speciation is advanced among the lineages from the Latin American clade and that these lineages fulfill the criteria to be considered phylogenetic species.

**FIGURE 2.**
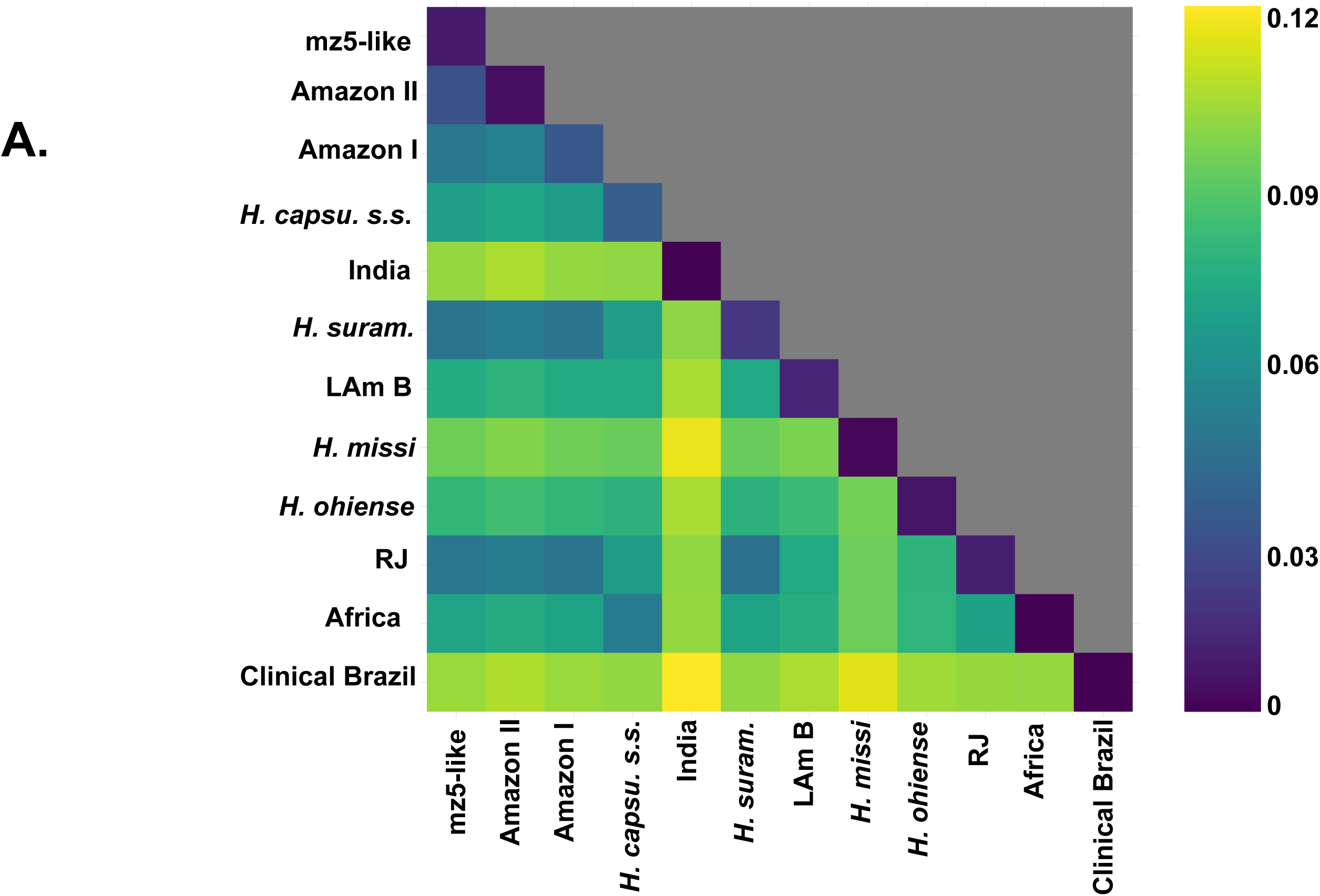
Genetic variation within and between lineages of *Histoplasma*. Interspecific genetic distance (Dxy) is larger than intraspecific variation (π) in the majority of pairwise comparisons. The upper diagonal shows π; all other squares show the pairwise distance between Dxy. Table S3 shows Fisher-Pittman permutation tests comparing the values of Dxy and π in all possible pairwise comparisons.

**FIGURE 3.**
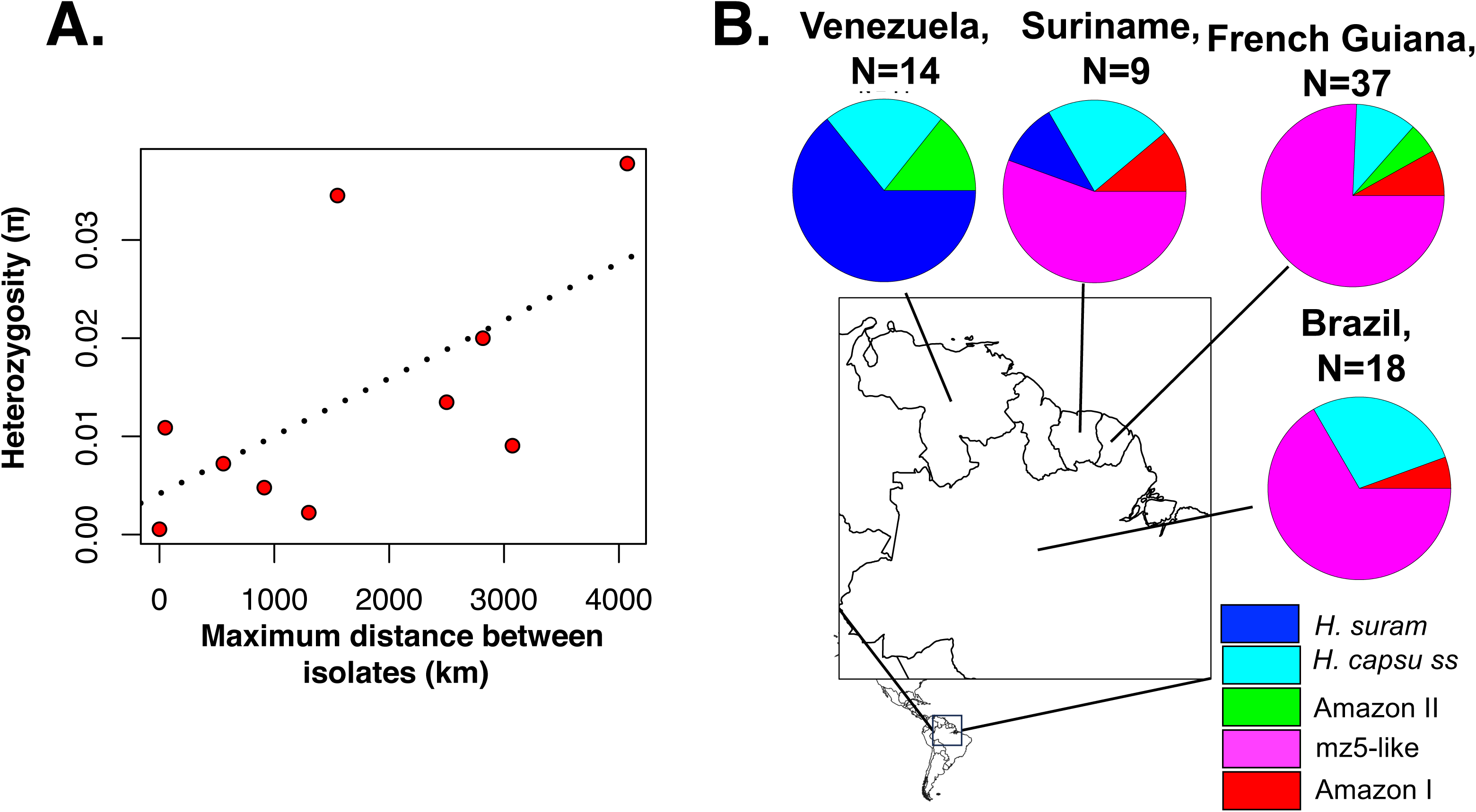
Genetic variation within and between lineages of *Histoplasma*. **A.** π, a metric of within-species genetic variation, is correlated with the geographic range size of each lineage, measured as the distance between the most distant isolates in a lineage. **B.** Proportional representation of the phylogenetic species identified in our sample in four countries of the Amazon basin.

We then compared some epidemiological aspects of histoplasmosis caused by each phylogenetic species. Out of the four countries that had at least 10 samples, three of them (Suriname, French Guiana, and Brazil) all showed a high prevalence of the mz5-like lineage (Figure 3B). Venezuela on the other hand showed a high prevalence of *H. suramericanum* and no mz5-like isolates. Consequently, the species composition of the Venezuela *Histoplasma* sample is the only that differs from other countries (X^2^ > 6.941, df = 1, *P* < 0.008, P_adjusted_ < 0.034). It is of note that isolates collected from Spain (N=4) were isolated from patients that migrated from South and Central America and all belong to *H. suramericanum* (Table S1).

Next, we studied whether the species composition of the sample changed from year to year. Figure 4A shows the proportional representation of each lineage per year. The year with the highest numbers of cases was 2015 with 13 cases, while the lowest was 2011 with two cases. The mz5-like lineage accounted for over 50% of the cases reported in this study in all years except for 2009 in which it accounted for 42.9% of the samples. None of these proportions differed significantly from each other (2-sample test for equality of proportions with continuity correction: *X*^2^ = 1.641, df = 1, P = 0.200).

**FIGURE 4.**
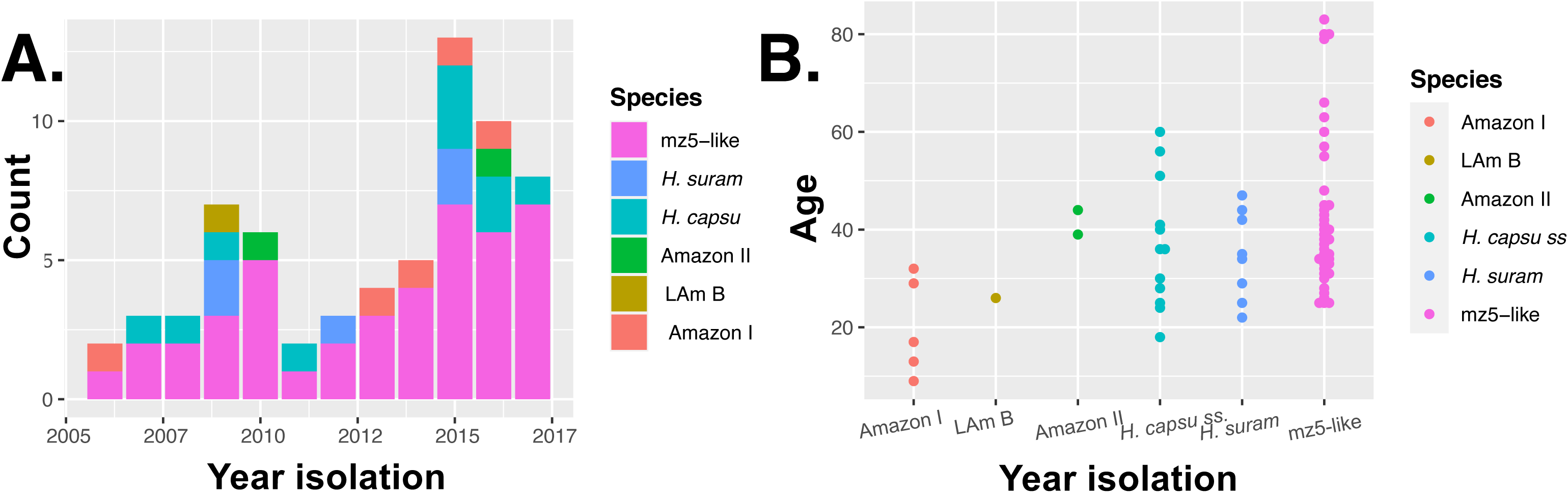
Epidemiological characteristics of the six *Histoplasma* phylogenetic species detected in the Amazon basin. **A.** Yearly proportions of each phylogenetic species in the sample presented here. **B.** Age distribution of the patients with histoplasmosis caused by each of the six phylogenetic species.

Third, we compared the mean patient age for each of the lineages identified. We detected significant differences among the age of patients affected by the five lineages (One-way ANOVA: F_5,62_ = 2.621, P = 0.032). Figure 4B shows that histoplasmosis patients caused by four lineages had a median age of ∼40 years old. Two lineages were significantly different from the others. Amazon I shows a younger median patient age (17.0 ± 10.05 years). mz5-like patients show a median older age (38.0 ± 16.24 years). Amazon II also has a high median age (41.5 years) but a low sample size (N = 2). Table S6 shows the Tukey HSD pairwise comparisons among lineages.

Finally, we compared the histoplasmosis sex ratio for each of the six phylogenetic species. Previous studies have noted that histoplasmosis is more common in men (48). All lineages show a higher proportional representation of male than female patients (Table 1). The mz5-like and *H. capsulatum sensu stricto* lineages show sufficient power for the comparison (Table 1). The mz5-like lineage shows no significant differences in the frequency of male and female patients. *Histoplasma capsulatum ss.*, on the other hand, was almost exclusively isolated from male patients (Table 1). Table S7 lists the linear regression coefficients indicating that *H. capsulatum ss* seems to have a higher male:female patient sex ratio than the other species present in the Amazon basin.

**TABLE 1.** Number of cases of histoplasmosis caused by each lineage in the Amazon basin. We also report the number of patients who are HIV+, of patients with the disseminated form of the disease, and the sex ratio. Table S5 shows the Tukey HSD comparisons.

| Lineage | N | HIV+ | Disseminated | Sex | | | $\chi^2$ power | P-value |
| --- | --- | --- | --- | --- | --- | --- | --- | --- |
| | | | | Male | Female | Ratio | | $\chi^2$ test |
| mz5-like | 45 | 40 | 25 | 30 | 15 | 2.00 | 0.918 | $\chi^2 = 1.931$, df = 1, P = 0.165 |
| *H. capsulatum* ss. | 15 | 11 | 8 | 14 | 1 | 14.00 | 0.491 | $\chi^2 = 4.9658$, df = 1, P = 0.026 |
| Amazon-I | 8 | 4 | 3 | 5 | 3 | 1.67 | 0.293 | NA |
| Amazon-II | 4 | 4 | 2 | 2 | 2 | 2.00 | 0.232 | NA |
| *H. suram* | 15 | 3 | 2 | 6 | 2 | 3.00 | 0.293 | NA |
| LAm B | 1 | 1 | 0 | 0 | 1 | NA | NA | NA |

## DISCUSSION

*Histoplasma* is one of the most important fungal pathogens in the world and the disease burden of histoplasmosis is a major concern for public health. Genome sequencing has revealed the existence of multiple cryptic species within the genus (11,14,24). The high incidence of histoplasmosis, and the high rates of histoplasmin skin reactivity in South America have led to the hypothesis that *Histoplasma* harbors a high level of genetic diversity in this continent. A potential corollary is that such genetic diversity also has clinical implications for patients with histoplasmosis. In this report, we identify a monophyletic group that contains four previously unidentified and highly differentiated phylogenetic species endemic to South and Central America, all related to *H. suramericanum*, and show that South America is a biodiversity hotspot for *Histoplasma* that houses at least seven phylogenetic species. The second most diverse continent, to date, is North American with three phylogenetic species (49).

Our sampling also allowed us to show that the epidemiological trends of histoplasmosis differ depending on the lineage that cases the disease. We report two potentially epidemiologically important differences between lineages of *Histoplasma*, the age of the patients, and the sex ratio of the affected patient. Histoplasmosis patients are more frequently males and the ratio has been reported to be close to 3:1 (14,50). Our findings suggest differences between the *Histoplasma* species in the Amazon basin and the sex ratio varies between 3:2 (in Amazon I) to 14:1 (*H. capsulatum* sensu stricto). Nonetheless, these assessments need to be taken carefully. The epidemic drivers of the HIV epidemic may vary between territories leading to different age distributions and sex ratios. Furthermore, women are usually tested earlier for HIV and are more likely to seek medical care than men. By contrast outdoor physical labor and exposure to *H. capsulatum* may be more frequent among men. Controlled animal infections will be the ultimate test of whether different lineages of *Histoplasma* represent a different health risk for difference sexes.

The identification of phylogenetic species is just the first step to understand the genetic diversity of *Histoplasma*. Perhaps the most important question of all, whether different species are associated with differences in virulence and clinical presentation of histoplasmosis, remains largely understudied. In this study, we report epidemiological differences among lineages in South America but only a comparative assessment from multiple isolates from each *Histoplasma* lineage will reveal whether genetic differences among lineages also lead to phenotypic differences in traits of clinical importance. Future research should address whether different clades differ in virulence which in turn will address whether the epidemiological patterns observed in the cohort here reported are caused by genetic changes in each of the *Histoplasma* lineages.

Our study has caveats that should be noted. Even though South America harbors the highest number of phylogenetic species known in *Histoplasma* to date, our sampling does not allow us to definitively affirm that South America is the most diverse hotspot of *Histoplasma* diversity because the sampling in other continents has been scant. We refrain from naming additional species in this effort as it is highly likely that there are other groups around the world and taxonomy should not be fully revisited until a more global portrait emerges.

Clinical and epidemiological comparisons between different lineages of *Histoplasma* remain rare (Table S8, but see 14) but are an important frontier that can reveal the tempo and mode of evolution of virulence strategies in fungal pathogens. Overall, our results suggest that South America serves as a geographic reservoir of genetic diversity of *Histoplasma* and underscores the need for systematic collection of the agents of endemic mycoses across tropical regions to better understand their evolutionary history. Now that genome sequencing is at hand for most species, it should be fully deployed to understand the evolutionary history and epidemiological patterns of histoplasmosis and other endemic mycoses.

## Data availability and sharing

All sequenced *Histoplasma* genomes were deposited at Short Read Archive (SRA – See Supplementary Material) and alignment files (.bam) files were deposited at Dryad. Analytical code will be permanently deposited in Zenodo.

## Acknowledgements

We would like to thank our reviewers and members of the Matute lab for helpful comments on earlier versions of the manuscript. Anastasia Litvintseva and Lalitha Gade (CDC) donated strains, and we are grateful to them. This project used isolates issued from the EDIRAPHIS study and the Technical Cooperation Among Countries project entitled “Control of Histoplasmosis on HIV infected patients in the Guiana Shield” granted in French Guiana and Suriname by the European Regional Development Fund (N°PRESAGE: 31362), the Pan American Health Organization - World Health Organization through, the Agence Nationale de Recherche sur le Sida et les hépatites virales – Agence autonome de l’Inserm (ANRS project number12260), respectively. The present project on molecular epidemiology was granted by the Ministère des Outre Mers (convention 2014-324-0008). MMT is supported through funding from Fundação de Apoio à Pesquisa do Distrito Federal (FAPDF) - under award 00193-00001871/2023-99. VES, QZ and DRM were supported by the National Institute of Allergy and Infectious Disease of the National Institutes of Health (NIH) under Award R01AI153523 to DRM. The funders had no role in study design, data collection and analysis, decision to publish, or preparation of the manuscript.

## Ethics statement

Ethical approval was obtained by the Comité de Protection des Personnes (CPP2012-47) and the Commission Nationale Informatique et Libertés (CNIL913511). Biological collection for sample collection was approved (DC-2013-1902).

## First authors’ biography

Dr. Tani Ly is a recently minted Ph.D. from the Université de Guyane. She is interested in genomics, and epidemiology of neglected tropical diseases.

---

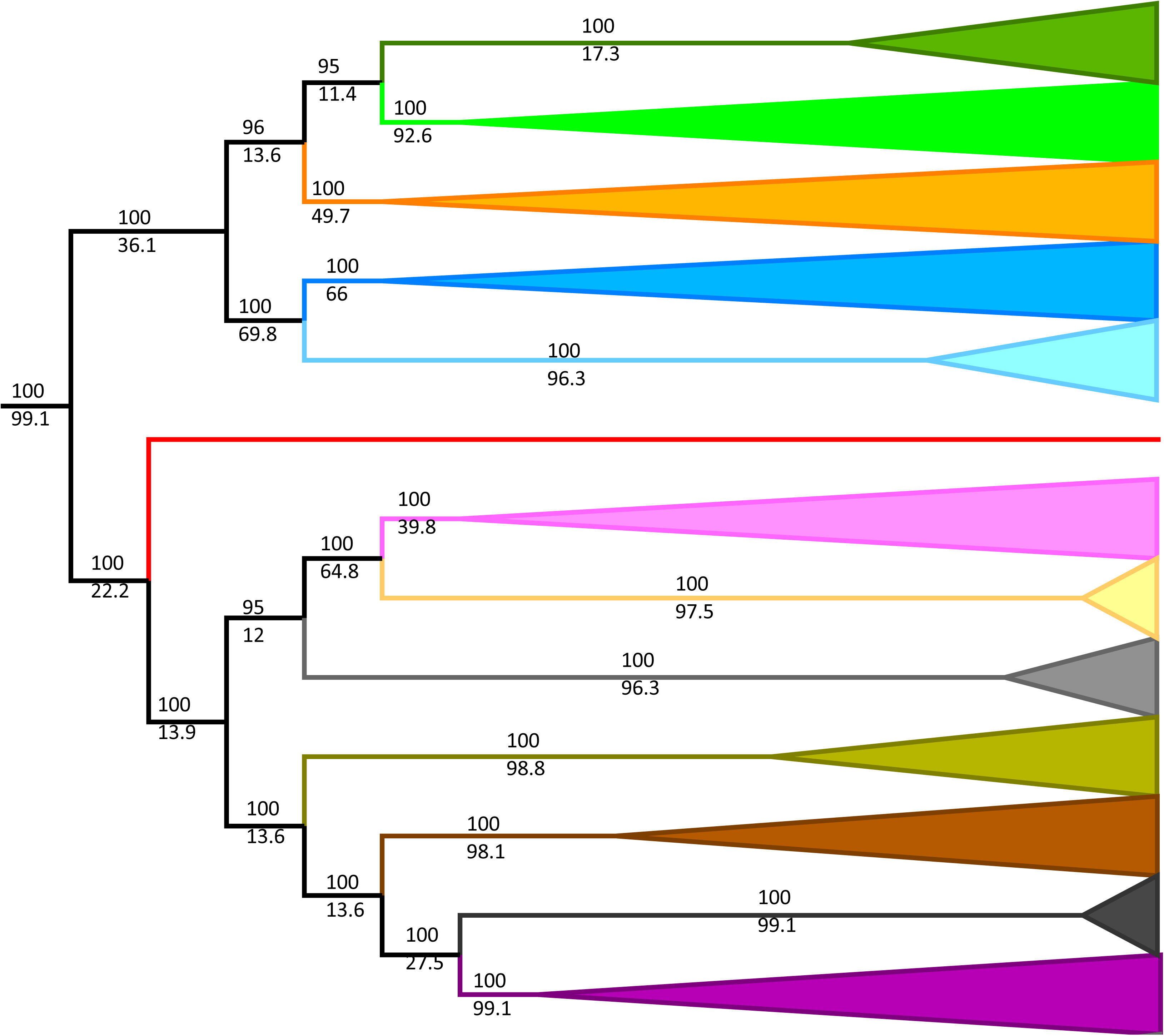

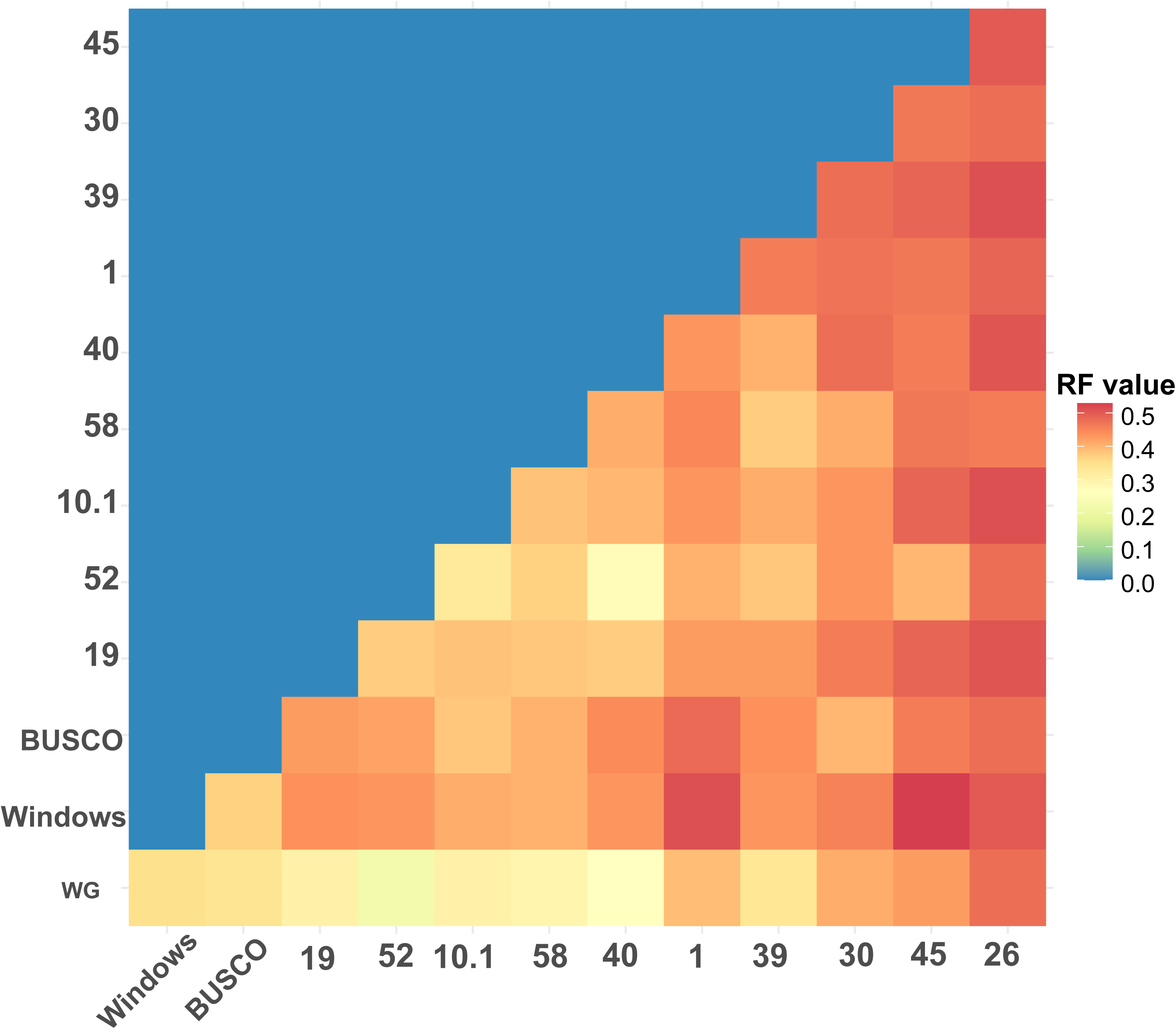

