## Supplementary Methods, Supplementary Tables, Supplementary Figure Legends for "High genetic diversity of *Histoplasma* in the Amazon basin, 2006-2017"

**SUPPLEMENTARY MATERIAL: High genetic diversity of *Histoplasma* in the Amazon basin, 2006-2017**

**SUPPLEMENTARY METHODS APPENDIX**

**DNA extraction**

After subculturing each isolate, we collected ~500 mg of the filamentous phase of *Histoplasma* spp. and heated the tissue at 65°C for 10 min. To extract DNA, we used mechanical lysis by adding 0.5 mm diameter zirconia/silica beads and lysis buffer to the preheated tissue and vigorously agitated the mixture using a Qiagen’s Vortex for 15 minutes. To purify DNA, we used the DNAeasy Blood and Tissue Kit (Qiagen, Hilden, Germany) according to the manufactures standard protocol. Finally, we evaluated DNA integrity and concentration using agarose-gel electrophoresis (0.8% w/v) and Qubit fluorimetry (Qubit fluorimeter, Thermo Fisher Scientific, Waltham, MA), respectively.

**SNP Variant calling**

First, we removed the Illumina adapters from all Illumina reads using Trimmomatic v 0.36 (A1). Then, we aligned trimmed reads to the *H. suramericanum MZ5* reference genome using BWA mem - v 0.7.15 (A2). Next, we identify single nucleotide polymorphisms (SNPs) using GATK 4.1.7.0 (A3, A4). We used the function HaplotypeCaller setting the -ploidy option as 1 (haploid). We then merged the resulting gvcf files using the GATK GenomicsDBImport function, and jointly genotyped the database with the GATK GenotypeGVCFs function. Finally, we filtered the multisample VCF file using GATK VariantFiltration with the following parameters: QD = 2.0 || FS_filter = 60.0 || MQ_filter = 30.0 || MQ_Rank_Sum_filter = -12.5 || Read_Pos_Rank_Sum_filter = -8.

**SUPPLEMENTARY FIGURE LEGENDS**

**FIGURE S1. Branch support for each monophyletic group calculated with the 100kb genomic windows.** Numbers on top of the branches are bootstrap support values; numbers under the branches are concordance factors. Lineages follow the same color scheme as Figures 1 and 2

**FIGURE S2.** **Robertson-Foulds (RF) distance between the topologies generated from different datasets, either each supercontig or the concatenated alignment.**

**SUPPLEMENTARY TABLES**

**TABLE S1.** List of *Histoplasma sp.* isolates used in the study

| Isolate Name | Clade | Country of isolation | Source | Sex | Country of birth | Nature of sampling | Disseminated /localized | Year of isolation |
| --- | --- | --- | --- | --- | --- | --- | --- | --- |
| FG-pia2052 | mz5-like | French Guiana | Human | f | French Guiana | arthritis | arthritis | 2009 |
| FG-pir2086 | mz5-like | French Guiana | Human | m |  | tongue | disseminated | 2009 |
| FG-ama2041 | mz5-like | French Guiana | Human/HIV+ | m | Guyana | bone marrow |  | 2010 |
| FG-bel2002 | mz5-like | French Guiana | Human/HIV+ | m | Haiti | blood | disseminated | 2017 |
| FG-bik2051 | mz5-like | French Guiana | Human/HIV+ | f | French Guiana | ganglion | disseminated | 2017 |
| FG-bon2001 | mz5-like | French Guiana | Human/HIV+ | m | Suriname | bone marrow | disseminated | 2015 |
| FG-cle2015 | mz5-like | French Guiana | Human/HIV+ | m | Haiti | liver | disseminated | 2012 |
| FG-cri2041 | mz5-like | French Guiana | Human/HIV+ | m | Saint-Lucia | Stomach | digestive | 2010 |
| FG-das2062 | mz5-like | French Guiana | Human/HIV+ | f | Brazil | colon | disseminated | 2013 |
| FG-dee2016 | mz5-like | French Guiana | Human/HIV+ | f | Suriname | bone marrow | digestive | 2010 |
| FG-deo2088 | mz5-like | French Guiana | Human/HIV+ | f | Brazil | cerebrospinal fluid | disseminated | 2014 |
| FG-fan2059 | mz5-like | French Guiana | Human/HIV+ | m | Suriname | Lymph node | ganglionic and colic | 2017 |
| FG-fer2036 | mz5-like | French Guiana | Human/HIV+ | m | Brazil | bone marrow | disseminated | 2017 |
| FG-gre 2022 | mz5-like | French Guiana | Human/HIV+ | f | Haiti | blood | disseminated | 2016 |
| FG-jos2044 | mz5-like | French Guiana | Human/HIV+ | m | French Guiana | colon | disseminated | 2016 |
| FG-kou2024 | mz5-like | French Guiana | Human/HIV+ | m | - | blood |  | 2017 |
| FG-lin2055 | mz5-like | French Guiana | Human/HIV+ | m | French Guiana | Bronchoalveolar lavage |  | 2014 |
| FG-mel2036 | mz5-like | French Guiana | Human/HIV+ | f | Brazil | liver | hepatic | 2007 |
| FG-non2028 | mz5-like | French Guiana | Human/HIV+ | m | Brazil | urine | disseminated | 2017 |
| FG-pie2055 | mz5-like | French Guiana | Human/HIV+ | m |  | bone marrow |  | 2017 |
| FG-pin2043 | mz5-like | French Guiana | Human/HIV+ | m |  | colon | digestive | 2010 |
| FG-poe2043 | mz5-like | French Guiana | Human/HIV+ | f |  | colon |  | 2012 |
| FG-rod2046 | mz5-like | French Guiana | Human/HIV+ | m | Brazil | bone marrow | disseminated | 2010 |
| FG-sou0318 | mz5-like | French Guiana | Human/HIV+ | m | Brazil | blood | disseminated | 2006 |
| FG-ver2032 | mz5-like | French Guiana | Human/HIV+ | f | Suriname | ganglion |  |  |
|  |  |  |  |  |  |  |  | 2009 |
| FG-zaa2004 | mz5-like | French Guiana | Human/HIV+ | f | Suriname | bone marrow | disseminated | 2007 |
| FG-zul2036 | mz5-like | French Guiana | Human/HIV+ | f | French Guiana | subclavicule | disseminated | 2009 |
| S-ada2068 | mz5-like | Suriname | Human/HIV+ | f | Suriname | bone marrow |  |  |
|  |  |  |  |  |  |  |  | 2014 |
| S-asa2073 | mz5-like | Suriname | Human/HIV+ | m | Suriname | bone marrow |  | 2014 |
| S-asaazp1 | mz5-like | Suriname | Human/HIV+ | m | Suriname | blood |  | 2013 |
| S-mis2065 | mz5-like | Suriname | Human/HIV+ | m | Suriname | bone marrow |  | 2014 |
| S-spa2057 | mz5-like | Suriname | Human/HIV+ | f | Suriname | liver |  | 2015 |
| FG-ada2079 | mz5-like | French Guiana | Human/HIV+ | m | French Guiana | bone marrow | disseminated | 2013 |
| HCAM | Amazon_II | Venezuela | Human/HIV+ | f | Venezuela | bone marrow | disseminated |  |
| HCM-H | Amazon_II | Venezuela | Human/HIV+ | m | Venezuela | - | disseminated |  |
| FG-bre2013 | Amazon_II | French Guiana | Human/HIV+ | m | Saint-Lucia | bone marrow |  | 2010 |
| FG-pic2055 | Amazon_II | French Guiana | Human/HIV+ | f | French Guiana | ganglion | ganglionic | 2016 |
| FG-aco2005 | Amazon_III | French Guiana | Human | m | French Guiana | blood | disseminated | 2013 |
| FG-bru | Amazon_III | French Guiana | Human/HIV+ | f | Cayenne | blood |  | 2006 |
| FG-pers2034 | Amazon_III | French Guiana | Human/HIV+ | m | French Guiana | bone marrow | disseminated | 2016 |
| S-dor2042 | Amazon_III | Suriname | Human/HIV+ | f | Suriname | - |  | 2014 |
| S-dij2058 | LAmA | Suriname | Human/HIV+ | m | Suriname | bone marrow |  | 2015 |
| HC7072a | LAmA | Venezuela | Human |  | Venezuela |  |  |  |
| HC7090 | LAmA | Venezuela | Human/HIV+ | m | Venezuela | skin |  |  |
| HC970591 | LAmA | Venezuela | Human/HIV+ | f | Venezuela | lung |  |  |
| hcjb | LAmA | Venezuela | Human | m | Venezuela |  | disseminated |  |
| HC1070058-2 | LAmA | Venezuela | Human | m | Venezuela |  | disseminated |  |
| HC4137 | LAmA | Venezuela | Human | f | Venezuela | skin |  |  |
| HC3066 | LAmA | Venezuela | Human |  |  |  |  |  |
| HC776 | LAmA | Venezuela | Human | m | Venezuela | erythematous plaques |  |  |
| HC394 | LAmA | Venezuela | Human | m | Venezuela | erythematous plaques |  |  |
| B06379 | LAmA | Nicaragua | Soil |  | - | - | - |  |
| CM5679 | LAmA | Spain | Human |  | Mexico |  |  | 2009 |
| CM5692 | LAmA | Spain | Human |  | Mexico |  |  | 2009 |
| CM7057 | LAmA | Spain | Human |  | Mexico |  |  | 2012 |
| CM7717 | LAmA | Spain | Human |  | Mexico |  |  | 2015 |
| A-290302130 | LAmB | Martinique | Human/HIV+ ? | f | Martinique | Bronchial aspiration |  | 2009 |
| HCR-P | Capsu | Venezuela | Human | m | Venezuela |  |  |  |
| HC3645 | Capsu | Venezuela | Human/HIV+ | m |  | Nasal granuloma |  |  |
| HC970588 | Capsu | Venezuela | Human | m | Venezuela |  | disseminated |  |
| FG-sat2037 | Capsu | French Guiana | Human | m |  | ascites | disseminated | 2009 |
| FG-dos0487 | Capsu | French Guiana | Human/HIV+ | m |  | bone marrow |  | 2007 |
| FG-martin | Capsu | French Guiana | Human/HIV+ | m | Brazil | blood | disseminated | 2017 |
| FG-wil2021 | Capsu | French Guiana | Human/HIV+ | m | French Guiana | blood | disseminated | 2008 |
| G-JJ | Capsu | Republic of Guyana | Human/HIV+ | m | Republic of Guyana | bone marrow | disseminated | 2015 |
| S-rob2039 | Capsu | Suriname | Human/HIV+ | m | Suriname | blood |  |  |
|  |  |  |  |  |  |  |  | 2014 |
| S-tan2075 | Capsu | Suriname | Human/HIV+ | m | Suriname | blood |  |  |
|  |  |  |  |  |  |  |  | 2014 |
| 11571 (Belem 1) | mz5-like | Brazil | Human | M | Brazil | ganglion |  | 2008 |
| 4363 (Belem 10) | mz5-like | Brazil | Human/HIV+ | F | Brazil | bone marrow | Disseminated | 2015 |
| 4107 (Belem 11) | mz5-like | Brazil | Human/HIV+ | M | Brazil | bone marrow | Disseminated | 2015 |
| 4516 (Belem 12) | Amazon_III | Brazil | Human/HIV+ | M | Brazil | bone marrow | Disseminated | 2015 |
| 4809 (Belem 13) | Capsu | Brazil | Human/HIV+ | M | Brazil | bone marrow |  | 2016 |
| 47692 (Belem 14) | mz5-like | Brazil | Human/HIV+ | M | Brazil | Urine | Disseminated | 2016 |
| 4265 (Belem 15) | mz5-like | Brazil | Human/HIV+ | M | Brazil | bone marrow | Disseminated | 2015 |
| 4729 (Belem 16) | mz5-like | Brazil | Human/HIV+ | M | Brazil | Urine | Disseminated | 2016 |
| 4337 (Belem 17) | mz5-like | Brazil | Human/HIV+ | M | Brazil | bone marrow | Disseminated | 2015 |
| 5215 (Belem 18) | mz5-like | Brazil | Human/HIV+ | M | Brazil | bone marrow | Disseminated | 2016 |
| 52292 (Belem 19) | Capsu | Brazil | Human/HIV+ | M | Brazil | bone marrow | Disseminated | 2016 |
| 11572 (Belem 2) | mz5-like | Brazil | Human | M | Brazil | Ganglion |  | 2008 |
| 5254 (Belem 20) | mz5-like | Brazil | Human/HIV+ | F | Brazil | bone marrow | Disseminated | 2016 |
| 2202 (Belem 5) | mz5-like | Brazil | Human/Transplant | M | Brazil | Bronchial aspiration |  | 2011 |
| 2353 (Belem 6) | Capsu | Brazil | Human | M | Brazil | Lung biopsy | Pulmonary | 2011 |
| 3948 (Belem 7) | mz5-like | Brazil | Human/HIV+ | M | Brazil | bone marrow | Disseminated | 2015 |
| 3865 (Belem 8) | Capsu | Brazil | Human/HIV+ | F | Brazil | bone marrow | Disseminated | 2015 |
| 4182 (Belem 9) | Capsu | Brazil | Human/HIV+ | M | Brazil | bone marrow | Disseminated | 2015 |

**TABLE S2.** SRA for sequences generated in this study.

| **Isolate Name** | **SRA Accession number** |
| --- | --- |
| A-290302130 | SRR31893250 |
| B06379 | SRR31893266 |
| CM5679 | SRR31893265 |
| CM5692 | SRR31893264 |
| CM7057 | SRR31893263 |
| CM7717 | SRR31893262 |
| FG-aco2005 | SRR31893282 |
| FG-ada2079 | SRR31893313 |
| FG-ama2041 | SRR31893312 |
| FG-bel2002 | SRR31893301 |
| FG-bik2051 | SRR31893289 |
| FG-bon2001 | SRR31893279 |
| FG-bre2013 | SRR31893285 |
| FG-bru0291 | SRR31893281 |
| FG-cle2015 | SRR31893268 |
| FG-cri2041 | SRR31893257 |
| FG-das2062 | SRR31893249 |
| FG-dee2016 | SRR31893248 |
| FG-deo2088 | SRR31893247 |
| FG-dos0487 | SRR31893277 |
| FG-fan2059 | SRR31893311 |
| FG-fer2036 | SRR31893310 |
| FG-gre 2022 | SRR31893309 |
| FG-jos2044 | SRR31893308 |
| FG-kou2024 | SRR31893307 |
| FG-lin2055 | SRR31893306 |
| FG-martin | SRR31893276 |
| FG-mel2036 | SRR31893305 |
| FG-non2028 | SRR31893304 |
| FG-pers2034 | SRR31893280 |
| FG-pia2052 | SRR31893303 |
| FG-pic2055 | SRR31893284 |
| FG-pie2055 | SRR31893302 |
| FG-pin2043 | SRR31893300 |
| FG-pir2086 | SRR31893299 |
| FG-poe2043 | SRR31893298 |
| FG-rod2046 | SRR31893296 |
| FG-sat2037 | SRR31893275 |
| FG-sou0318 | SRR31893295 |
| FG-ver2032 | SRR31893293 |
| FG-wil2021 | SRR31893274 |
| FG-zaa2004 | SRR31893286 |
| FG-zul2036 | SRR31893292 |
| G-JJ | SRR31893273 |
| HC1070058-2 | SRR31893258 |
| HC3066 | SRR31893256 |
| HC3645 | SRR31893270 |
| HC394 | SRR31893255 |
| HC4137 | SRR31893254 |
| HC7072a | SRR31893261 |
| HC7090 | SRR31893260 |
| HC776 | SRR31893253 |
| HC970588 | SRR31893269 |
| HC970591 | SRR31893252 |
| HCAM | SRR31893287 |
| hcjb | SRR31893251 |
| HCM-H | SRR31893283 |
| HCR-P | SRR31893267 |
| S-ada2068 | SRR31893291 |
| S-asa2073 | SRR31893297 |
| S-asaazp1 | SRR31893290 |
| S-dij2058 | SRR31893259 |
| S-dor2042 | SRR31893278 |
| S-mis2065 | SRR31893294 |
| S-rob2039 | SRR31893272 |
| S-spa2057 | SRR31893288 |
| S-tan2075 | SRR31893271 |

**TABLE S3.** SRA accession numbers for accession previously sequenced and used in this study.

| **Isolate name** | **SRA accession number** | **Phylogenetic species** | **Reference** |
| --- | --- | --- | --- |
| 104_p_06_S19 | SRR27481878 | India | (A5) |
| 104_P_19_S5 | SRR27481877 | India | (A5) |
| 107_P_06_S1 | SRR27481870 | India | (A5) |
| 117_p_12_S17 | SRR27481869 | India | (A5) |
| 122_p_10_B_S15 | SRR27481868 | India | (A5) |
| 136_P_07_S6 | SRR27481867 | India | (A5) |
| 144_p_08_S14 | SRR27481866 | India | (A5) |
| 1517_p_17_S20 | SRR27481865 | India | (A5) |
| 256_P_18_S2 | SRR27481864 | India | (A5) |
| 316_p_10_S18 | SRR27481863 | India | (A5) |
| 327_P_12_S7 | SRR27481876 | India | (A5) |
| 343_p_18_S1 | SRR27481875 | India | (A5) |
| 388_p_11_S16 | SRR27481874 | India | (A5) |
| S11-105_p_06 | SRR27481873 | India | (A5) |
| S14-108_p_06 | SRR27481872 | India | (A5) |
| S16-106_p_06 | SRR27481871 | India | (A5) |
| 4 | SRR8084704 | RJ | (A6) |
| 3 | SRR8084705 | RJ | (A6) |
| 2 | SRR8084706 | RJ | (A6) |
| 1 | SRR8084707 | RJ | (A6) |
| 8 | SRR8084708 | RJ | (A6) |
| 7 | SRR8084709 | RJ | (A6) |
| 6 | SRR8084710 | RJ | (A6) |
| 12 | SRR8084714 | RJ | (A6) |
| 11 | SRR8084715 | RJ | (A6) |
| 14 | SRR8084716 | RJ | (A6) |
| 13 | SRR8084717 | RJ | (A6) |
| 15 | SRR8084719 | RJ | (A6) |
| 18 | SRR8084720 | RJ | (A6) |
| 17 | SRR8084721 | RJ | (A6) |
| G222B | SRX3350818 | *H. ohiense* | (A7) |
| G217B | SRX3350817 | *H. ohiense* | (A7) |
| CI_10 | SRX3350821 | *H. ohiense* | (A7) |
| CI_4 | SRX3350841 | *H. ohiense* | (A7) |
| CI_17 | SRX3350822 | *H. ohiense* | (A7) |
| CI_9 | SRX3350820 | *H. ohiense* | (A7) |
| CI_30 | SRX3350824 | *H. ohiense* | (A7) |
| CI_18 | SRX3350823 | *H. ohiense* | (A7) |
| CI_6 | SRX3350819 | *H. ohiense* | (A7) |
| CI_35 | SRX3350825 | *H. ohiense* | (A7) |
| CI_24 | SRX3350845 | *H. mississipiense* | (A7) |
| CI_43 | SRX3350837 | *H. mississipiense* | (A7) |
| CI_22 | SRX3350842 | *H. mississipiense* | (A7) |
| CI_7 | SRX3350840 | *H. mississipiense* | (A7) |
| CI_42 | SRX3350844 | *H. mississipiense* | (A7) |
| 505 | SRX3350830 | *H. mississipiense* | (A7) |
| DOWNS | SRX3350816 | *H. mississipiense* | (A7) |
| CI_19 | SRX3350843 | *H. mississipiense* | (A7) |
| WU24 | SRX3350838 | *H. mississipiense* | (A7) |
| UCLA_531 | SRX3350836 | *H. mississipiense* | (A7) |
| 21_14 | SRX3350835 | *H. suramericanum* | (A7) |
| 3_11G | SRX3350833 | *H. suramericanum* | (A7) |
| 27_14 | SRX3350832 | *H. suramericanum* | (A7) |
| 1986 | SRX3350827 | *H. capsulatum ss* | (A7) |
| MZ5 | SRX3350829 | *H. capsulatum ss* | (A7) |
| G186A | SRX3350828 | *H. capsulatum ss* | (A7) |
| G184A | SRX3350831 | *H. capsulatum ss* | (A7) |
| MV3 | SRX3350826 | N/D | (A7) |
| duboisii_A | SRX3350834 | Africa | (A7) |
| duboisii_B | SRX3350839 | Africa | (A7) |
| 109_P_06_S4 | PRJNA1201237 | *B. dermatitidis* | (A8) |
| 143_P_08_S8 | PRJNA1201237 | *B. dermatitidis* | (A8) |
| Dr_Anuradha_Fungal_WGS_S13 | PRJNA1201237 | *B. dermatitidis* | (A8) |
| Ep_130_s_7 | PRJNA1201237 | *B. parvus* | (A9) |
| ep139_s_1 | PRJNA178178 | *B. silverae* (previously classified as *B. parvus*) | (A9) |
| Pb_339 | SRR4024750 | *P. restrepiensis* | (A10) |
| Pb_60855 | SRR4024748 | *P. restrepiensis* | (A10) |
| Pb_66_ATCACG_L001 | SAMN05171529 | *P. brasiliensis* | (A10) |
| Pb_jam | SRR4024745 | *P. restrepiensis* | (A10) |
| PbD02_TAGCTT_L001 | SRR4024744 | *P. brasiliensis* | (A10) |
| Ep_9510_s_8 | PRJNA416769 | *E. pasteurianus* | (A9) |
| Ec_4076_s_7 | PRJNA178252 | *E. crescens* | (A9) |
| s_2 | PRJNA178252 | *E. crescens* | (A9) |

**TABLE S4. Modeltest results showing the fit for a variety of molecular evolution models.**

| **Model** | **LogLik** | **df** | **AIC** | **AICc** | **BIC** |
| --- | --- | --- | --- | --- | --- |
| TVMe+R3 | 103668491 | 357 | 207337695 | 207337695 | 207342619 |
| SYM+R3 | 103668573 | 358 | 207337863 | 207337863 | 207342800 |
| TVM+F+R3 | 103670318 | 360 | 207341356 | 207341357 | 207346321 |
| GTR+F+R3 | 103670417 | 361 | 207341557 | 207341557 | 207346536 |
| GTR+F+R4 | 103670408 | 363 | 207341542 | 207341542 | 207346548 |
| TIM3e+R3 | 103689435 | 356 | 207379583 | 207379583 | 207384493 |
| TIM2e+R3 | 103690032 | 356 | 207380777 | 207380777 | 207385686 |
| TPM3+F+R3 | 103695571 | 358 | 207391858 | 207391859 | 207396796 |
| TPM3u+F+R3 | 103695571 | 358 | 207391858 | 207391859 | 207396796 |
| TIM3+F+R3 | 103695572 | 359 | 207391861 | 207391861 | 207396813 |
| K3P+R3 | 103696528 | 355 | 207393766 | 207393766 | 207398662 |
| TIMe+R3 | 103696527 | 356 | 207393765 | 207393765 | 207398675 |
| TPM2u+F+R3 | 103696580 | 358 | 207393877 | 207393877 | 207398814 |
| TPM2+F+R3 | 103696580 | 358 | 207393877 | 207393877 | 207398814 |
| TIM2+F+R3 | 103696589 | 359 | 207393897 | 207393897 | 207398848 |
| K2P+R3 | 103702989 | 354 | 207406685 | 207406685 | 207411568 |
| TNe+R3 | 103702987 | 355 | 207406685 | 207406685 | 207411581 |
| K3Pu+F+R3 | 103707149 | 358 | 207415014 | 207415014 | 207419951 |
| TIM+F+R3 | 103707149 | 359 | 207415016 | 207415016 | 207419967 |
| HKY+F+R3 | 103713799 | 357 | 207428311 | 207428311 | 207433235 |
| TN+F+R3 | 103713799 | 358 | 207428313 | 207428313 | 207433250 |
| GTR+F+R2 | 104383577 | 359 | 208767872 | 208767872 | 208772823 |
| GTR+F+G4 | 106154285 | 358 | 212309285 | 212309285 | 212314223 |
| GTR+F+I+G4 | 106154285 | 359 | 212309287 | 212309287 | 212314238 |
| GTR+F | 112489650 | 357 | 224980015 | 224980015 | 224984938 |
| GTR+F+I | 112489657 | 358 | 224980031 | 224980031 | 224984968 |
| JC+R3 | 116591713 | 353 | 233184131 | 233184131 | 233189000 |
| F81+F+R3 | 116614415 | 356 | 233229541 | 233229542 | 233234451 |

**TABLE S5**. Fisher Pittman permutation comparing values of intraspecific heterozygosity across *Histoplasma* species.

| **Species 1** | **Species 2** | **π̅Species1** | **π̅Species2** | **Dxy** | **Z value** | **P value** |
| --- | --- | --- | --- | --- | --- | --- |
| Amazon II | Amazon I | 0.00478 | 0.00905 | 0.03139 | -36.286 | < 0.0001 |
| Amazon III | Amazon I | 0.03453 | 0.00905 | 0.05077 | -38.201 | < 0.0001 |
| Amazon III | Amazon II | 0.03453 | 0.00478 | 0.05526 | -6.753 | < 0.0001 |
| *H. capsulatum* | Amazon I | 0.03779 | 0.00905 | 0.07106 | -47.530 | < 0.0001 |
| *H. capsulatum* | Amazon II | 0.03779 | 0.00478 | 0.07565 | -17.518 | < 0.0001 |
| *H. capsulatum* | Amazon III | 0.03779 | 0.03453 | 0.06946 | -19.785 | < 0.0001 |
| Clinical | Amazon I | 0.00014 | 0.00905 | 0.10753 | -34.894 | < 0.0001 |
| Clinical | Amazon II | 0.00014 | 0.00478 | 0.11182 | -4.470 | < 0.0001 |
| Clinical | Amazon III | 0.00014 | 0.03453 | 0.10742 | -6.340 | < 0.0001 |
| Clinical | *H. capsulatum* | 0.00014 | 0.03779 | 0.10603 | -16.492 | < 0.0001 |
| Clinical | India | 0.00014 | 0.00054 | 0.12647 | -13.038 | < 0.0001 |
| Clinical | LAm A | 0.00014 | 0.02000 | 0.10612 | -12.643 | < 0.0001 |
| Clinical | LAm B | 0.00014 | 0.01348 | 0.11052 | -2.990 | 0.00410 |
| Clinical | *H. mississippiense* | 0.00014 | 0.00223 | 0.12098 | -8.062 | < 0.0001 |
| Clinical | *H. ohiense* | 0.00014 | 0.00722 | 0.10857 | -8.768 | < 0.0001 |
| Clinical | Rio | 0.00014 | 0.01088 | 0.10695 | -12.250 | < 0.0001 |
| Clinical | 27-14 | 0.00014 | NA | 0.11063 | -1.414 | 0.33523 |
| Clinical | *H. capsulatum var. duboisii* | 0.00014 | 0.00020 | 0.10672 | -2.236 | 0.06961 |
| *H. capsulatum var. duboisii* | Amazon I | 0.00020 | 0.00905 | 0.07463 | -34.783 | < 0.0001 |
| *H. capsulatum var. duboisii* | Amazon II | 0.00020 | 0.00478 | 0.07908 | -4.468 | < 0.0001 |
| *H. capsulatum var. duboisii* | Amazon III | 0.00020 | 0.03453 | 0.07374 | -5.795 | < 0.0001 |
| *H. capsulatum var. duboisii* | *H. capsulatum* | 0.00020 | 0.03779 | 0.05283 | -10.268 | < 0.0001 |
| *H. capsulatum var. duboisii* | India | 0.00020 | 0.00054 | 0.10668 | -13.038 | < 0.0001 |
| *H. capsulatum var. duboisii* | LAm A | 0.00020 | 0.02000 | 0.07387 | -12.113 | < 0.0001 |
| *H. capsulatum var. duboisii* | LAm B | 0.00020 | 0.01348 | 0.07933 | -2.979 | 0.00460 |
| *H. capsulatum var. duboisii* | *H. mississippiense* | 0.00020 | 0.00223 | 0.09830 | -8.061 | < 0.0001 |
| *H. capsulatum var. duboisii* | *H. ohiense* | 0.00020 | 0.00722 | 0.08252 | -8.762 | < 0.0001 |
| *H. capsulatum var. duboisii* | Rio | 0.00020 | 0.01088 | 0.07301 | -12.146 | < 0.0001 |
| *H. capsulatum var. duboisii* | 27-14 | 0.00020 | NA | 0.07774 | -1.414 | 0.33043 |
| India | Amazon I | 0.00054 | 0.00905 | 0.10666 | -45.523 | < 0.0001 |
| India | Amazon II | 0.00054 | 0.00478 | 0.11112 | -15.163 | < 0.0001 |
| India | Amazon III | 0.00054 | 0.03453 | 0.10674 | -16.924 | < 0.0001 |
| India | *H. capsulatum* | 0.00054 | 0.03779 | 0.10588 | -26.423 | < 0.0001 |
| LAm A | Amazon I | 0.02000 | 0.00905 | 0.04883 | -44.606 | < 0.0001 |
| LAm A | Amazon II | 0.02000 | 0.00478 | 0.05255 | -13.595 | < 0.0001 |
| LAm A | Amazon III | 0.02000 | 0.03453 | 0.04810 | -14.389 | < 0.0001 |
| LAm A | *H. capsulatum* | 0.02000 | 0.03779 | 0.07045 | -25.531 | < 0.0001 |
| LAm A | India | 0.02000 | 0.00054 | 0.10524 | -23.280 | < 0.0001 |
| LAm B | Amazon I | 0.01348 | 0.00905 | 0.07746 | -35.558 | < 0.0001 |
| LAm B | Amazon II | 0.01348 | 0.00478 | 0.08173 | -5.179 | < 0.0001 |
| LAm B | Amazon III | 0.01348 | 0.03453 | 0.07690 | -6.702 | < 0.0001 |
| LAm B | *H. capsulatum* | 0.01348 | 0.03779 | 0.07663 | -16.293 | < 0.0001 |
| LAm B | India | 0.01348 | 0.00054 | 0.11009 | -13.738 | < 0.0001 |
| LAm B | LAm A | 0.01348 | 0.02000 | 0.07606 | -13.066 | < 0.0001 |
| *H. mississippiense* | Amazon I | 0.00223 | 0.00905 | 0.09872 | -40.593 | < 0.0001 |
| *H. mississippiense* | Amazon II | 0.00223 | 0.00478 | 0.10309 | -10.195 | < 0.0001 |
| *H. mississippiense* | Amazon III | 0.00223 | 0.03453 | 0.09885 | -11.829 | < 0.0001 |
| *H. mississippiense* | *H. capsulatum* | 0.00223 | 0.03779 | 0.09737 | -21.640 | < 0.0001 |
| *H. mississippiense* | India | 0.00223 | 0.00054 | 0.12312 | -18.707 | < 0.0001 |
| *H. mississippiense* | LAm A | 0.00223 | 0.02000 | 0.09736 | -18.326 | < 0.0001 |
| *H. mississippiense* | LAm B | 0.00223 | 0.01348 | 0.10176 | -8.764 | < 0.0001 |
| *H. ohiense* | Amazon I | 0.00722 | 0.00905 | 0.08335 | -41.301 | < 0.0001 |
| *H. ohiense* | Amazon II | 0.00722 | 0.00478 | 0.08770 | -10.900 | < 0.0001 |
| *H. ohiense* | Amazon III | 0.00722 | 0.03453 | 0.08321 | -12.436 | < 0.0001 |
| *H. ohiense* | *H. capsulatum* | 0.00722 | 0.03779 | 0.08156 | -21.764 | < 0.0001 |
| *H. ohiense* | India | 0.00722 | 0.00054 | 0.11017 | -19.398 | < 0.0001 |
| *H. ohiense* | LAm A | 0.00722 | 0.02000 | 0.08156 | -18.951 | < 0.0001 |
| *H. ohiense* | LAm B | 0.00722 | 0.01348 | 0.08617 | -9.473 | < 0.0001 |
| *H. ohiense* | *H. mississippiense* | 0.00722 | 0.00223 | 0.09995 | -14.443 | < 0.0001 |
| Rio | Amazon I | 0.01088 | 0.00905 | 0.04946 | -44.670 | < 0.0001 |
| Rio | Amazon II | 0.01088 | 0.00478 | 0.05339 | -14.207 | < 0.0001 |
| Rio | Amazon III | 0.01088 | 0.03453 | 0.04855 | -14.767 | < 0.0001 |
| Rio | *H. capsulatum* | 0.01088 | 0.03779 | 0.06933 | -24.148 | < 0.0001 |
| Rio | India | 0.01088 | 0.00054 | 0.10625 | -22.865 | < 0.0001 |
| Rio | LAm A | 0.01088 | 0.02000 | 0.04731 | -21.328 | < 0.0001 |
| Rio | LAm B | 0.01088 | 0.01348 | 0.07667 | -12.910 | < 0.0001 |
| Rio | *H. mississippiense* | 0.01088 | 0.00223 | 0.09826 | -17.922 | < 0.0001 |
| Rio | *H. ohiense* | 0.01088 | 0.00722 | 0.08248 | -18.634 | < 0.0001 |
| 27-14 | Amazon I | NA | 0.00905 | 0.07236 | -33.879 | < 0.0001 |
| 27-14 | Amazon II | NA | 0.00478 | 0.07637 | -3.738 | 0.00030 |
| 27-14 | Amazon III | NA | 0.03453 | 0.07133 | -4.858 | < 0.0001 |
| 27-14 | *H. capsulatum* | NA | 0.03779 | 0.07572 | -13.386 | < 0.0001 |
| 27-14 | India | NA | 0.00054 | 0.10949 | -12.329 | < 0.0001 |
| 27-14 | LAm A | NA | 0.02000 | 0.06717 | -10.551 | < 0.0001 |
| 27-14 | LAm B | NA | 0.01348 | 0.08205 | -2.235 | 0.10051 |
| 27-14 | *H. mississippiense* | NA | 0.00223 | 0.10127 | -7.347 | < 0.0001 |
| 27-14 | *H. ohiense* | NA | 0.00722 | 0.08541 | -8.047 | < 0.0001 |
| 27-14 | Rio | NA | 0.01088 | 0.07068 | -11.304 | < 0.0001 |

**TABLE S6. Pairwise comparisons reveal a difference in patient age among *Histoplasma* species in the Amazon basin. All comparisons were done with a post-hoc Tukey test (with multiple comparison corrections).** All t-tests were done with 62 degrees of freedom (i.e., the number of residual degrees of freedom in a One-way ANOVA).

| **Hypothesis** | **Estimate** | **Std. Error** | **t value** | **Pr(>\|t\|)** |
| --- | --- | --- | --- | --- |
| LAmB -  Amazon I == 0 | 6.000 | 15.986 | 0.375 | 0.9988 |
| Amazon II -  Amazon I == 0 | 21.500 | 12.210 | 1.761 | 0.4608 |
| *H. capsu* -  Amazon I == 0 | 17.083 | 7.768 | 2.199 | 0.2246 |
| LAmA -  Amazon I == 0 | 14.750 | 8.319 | 1.773 | 0.4532 |
| mz5-like -  Amazon I == 0 | 22.925 | 6.922 | 3.312 | 0.0155 * |
| AmazonII -  LAmB == 0 | 15.500 | 17.873 | 0.867 | 0.9445 |
| *H. capsu* -  LAmB == 0 | 11.083 | 15.189 | 0.730 | 0.9732 |
| *H. suram* -  LAmB == 0 | 8.750 | 15.478 | 0.565 | 0.9914 |
| mz5-like -  LAmB == 0 | 16.925 | 14.775 | 1.146 | 0.8400 |
| *H. capsu* - Amazon_II == 0 | -4.417 | 11.146 | -0.396 | 0.9984 |
| *H. suram* - Amazon_II == 0 | -6.750 | 11.537 | -0.585 | 0.9900 |
| mz5-like - Amazon_II == 0 | 1.425 | 10.574 | 0.135 | 1.0000 |
| *H. suram* - *H. capsu* == 0 | -2.333 | 6.661 | -0.350 | 0.9991 |
| mz5-like - *H. capsu* == 0 | 5.842 | 4.803 | 1.216 | 0.8039 |
| mz5-like - *H. suram* == 0 | 8.175 | 5.652 | 1.446 | 0.6662 |

**TABLE S7.** **Binomial regression coefficients comparing the patient sex ratio for the *Histoplasma* phylogenetic species present in the Amazon basin.**

|  | **Estimate** | **Std. Error** | **z value** | **Pr(>\|z\|)** |
| --- | --- | --- | --- | --- |
| **Intercept** | -1.995e-15 | 1.000 | 0.000 | 1.000 |
| **Amazon I** | -0.511 | 1.238 | -0.413 | 0.680 |
| ***H. capsulatum ss.*** | **2.639** | **1.439** | **-1.834** | **0.066** |
| **LAmB** | 0.156 | 1,455.00 | 0.011 | 0.992 |
| **mz5-like** | -0.693 | 1.049 | -0.661 | 0.509 |
| ***H. suramericanum*** | 0.154 | 1.144 | 0.135 | 0.893 |

**TABLE S8. Clinical and epidemiological data availability for the different phylogenetic species of *Histoplasma*. We only included studies that have used whole genome data.**

| **Study** | **Species included in the study** | **Clinical and epidemiological data available** |
| --- | --- | --- |
| (A7) | *H. ohiensis, H. mississippiensis, H. suramericanum*, *H. capsulatum sensu stricto*, Africa | None |
| (A5) | India | None |
| ^3^ | *H. ohiensis, H. mississippiensis, H. suramericanum* | None |
| (A6) | RJ | Yes, for the RJ lineage. |
